# Effects of mental imagery training on cognitive function and brain connectivity in people with Parkinson’s disease: A randomized pilot trial

**DOI:** 10.1101/2025.03.14.25324001

**Authors:** Jared Cherry, Alyssa M. Nelson, Lauren A. Robinson, Josh Goldstein, Ana Vives-Rodriguez, Emily Sharp, Sule Tinaz

## Abstract

Cognitive impairment is a debilitating problem in Parkinson’s disease (PD) with no effective treatment. We developed a personalized mental imagery (MI) intervention focusing on goal-directed activities and examined its effect on everyday cognitive functioning and brain functional connectivity in people with PD in a pilot randomized controlled trial (ClinicalTrials.gov identifier NCT05495997).

Thirty nondemented people with PD were randomized to PD-MI and PD-Control groups. During the six-week training period, PD-MI received MI training and PD-Control received psychoeducation on cognitive health in PD. Participants underwent cognitive and functional MRI assessments at baseline, six weeks, and 18 weeks. The primary outcomes included changes in Neuro-QoL Cognitive Function (CF) survey scores and functional connectivity.

The PD-MI compared to the PD-Control group showed (1) significant difference in Neuro-QoL-CF scores (F(1,26) = 6.802, p = 0.015) at six weeks which was not sustained at 18 weeks, (2) stronger connectivity between frontoparietal regions (T = 4.1, p = 0.009) during MI tasks at six weeks, and (3) weaker connectivity between visuospatial and motor regions at 18 weeks.

Personalized MI training can be effective in facilitating cognitive preparedness for everyday tasks in people with PD. Its long-term effects and feasibility in cognitively impaired PD cohorts need further investigation.

## Introduction

Cognitive impairment is a common and debilitating nonmotor symptoms of Parkinson’s disease (PD).^1^ The PD community identified maintaining cognitive function as one of its major unmet needs.^2^ Pharmacological treatment provides, at most, modest symptomatic benefit in PD-related dementia.^3^ Studies using lifestyle activities (e.g., exercise, mind-body interventions)^4^ and cognitive training programs^5^ in older adults and in people with PD have yielded promising but inconclusive evidence for cognitive benefits.^6^

Cognitive training programs in nondemented people with PD using structured paper-pencil materials,^7^ games,^8^ strategy training^9^, or computerized tasks^10^ have gained popularity with varying degrees of success for the cognitive domain that is treated (e.g., processing speed,^7^ working memory,^8^ and executive functions^9^), but usually without generalizable real-life benefits.^2^ The importance of transfer of training to relevant real-world tasks has also been highlighted by the Institute of Medicine’s criteria for these programs.^11^

In this study, we developed a cognitive training protocol based on mental imagery (MI) focusing on real-world tasks for people with PD. The protocol was meant to provide skill training in MI, so that participants could learn how to practice imagery to mentally prepare for everyday tasks. Mental imagery refers to the mental re-creation of perceptual experiences across sensory modalities, of past or future events and scenarios, and of motor acts with the potential to prepare the individual for action.^12^ It has been used extensively as a performance-enhancing strategy in sports and skilled performance,^13–16^ as a mnemonic strategy,^17,18^ and as an adjunct intervention in cognitive behavioral therapy for mood regulation.^19,20^ The embodied and experiential aspects of MI have been found to improve self-efficacy (e.g., to overcome fear of falling in older adults^21^), increase motivation, and promote engagement in planned activities and healthy behaviors.^22–24^ Mental imagery also plays a fundamental role in episodic future thinking, i.e., the ability to imagine the future that serves many cognitive functions (e.g., decision making, problem solving, emotion regulation, intention formation, planning) and is supported by the medial temporal memory and frontal-executive systems.^25^ Deficits in episodic future thinking have been shown in people with PD and attributed to executive dysfunction.^26^ Functional MRI (fMRI) studies of episodic future thinking including planning tasks (e.g., travel) and imagining everyday goals (e.g., clean out the room) showed activations in the default mode and frontoparietal executive networks.^27–31^

The MI training protocol was inspired by our previous work that demonstrated improvement in motor function in people with PD after kinesthetic motor imagery training. This improvement also correlated significantly with the functional connectivity strength between the right insula and dorsomedial frontal cortex, brain regions involved in motivating intentional action.^32^ Moreover, in a follow-up study, we demonstrated that people with PD with worse disease severity showed poorer vividness and richness of visual imagery content when they imagined static scenes (e.g., landscapes, indoor places), while the quality of visual imagery showed positive correlations with the functional connectivity of the visuospatial and sensorimotor networks.^33^

Here, we used MI to train the participants to simulate the planning and implementation process of personal goals in everyday life. Our primary objective was twofold: To examine the effects of the six-week MI training, compared to the psychoeducation control condition, on (1) daily cognitive functioning and (2) functional connectivity of the brain networks using fMRI. Given the personalized nature of MI training, we chose to use a self-reported measure as the primary outcome of daily cognitive functioning, namely, the Quality of Life in Neurological Disorders Cognitive Function survey (Neuro-QoL-CF)^34^ version 2, which measures the perceived difficulties in cognitive abilities and in the application of such abilities to everyday tasks. It provides valid, clinically meaningful, and generalizable assessments of cognitive functional abilities, has been validated in PD cohorts, and is intended for use in both clinical trials and clinical practice.

We hypothesized that MI training would result in (1) better daily cognitive functioning and (2) stronger functional connectivity between the brain networks involved in the imagery of goal-directed activities. Our secondary objective was to examine whether the hypothesized cognitive and brain effects of the MI training would be sustained over the course of 12 weeks.

## Materials and Methods

### Overview

This study was designed as a single-site pilot randomized controlled trial (NCT05495997) conducted at the Yale School of Medicine and Magnetic Resonance Research Center (see CONSORT checklist in Supplement 1). We assigned two groups randomly in parallel. The experimental PD-MI group received MI training, and the control PD-Con group received psychoeducation (Figure 1). In previous cognitive training trials, the training periods were anywhere between 4-12 weeks, training was usually administered in small groups at least once a week with some components administered in individual sessions, and usually combined with unsupervised practice (e.g., homework).^6^ Here, we chose the training and retention periods as six and 12 weeks, respectively. Building on our previous experience,^32^ we implemented a rigorous MI training protocol that was delivered during one-on-one sessions and practiced via homework. We also prepared psychoeducation modules and quizzes for the PD-Con group. All tests and fMRI scans were performed when participants were on their regular PD medication regimen.

**Figure 1.**
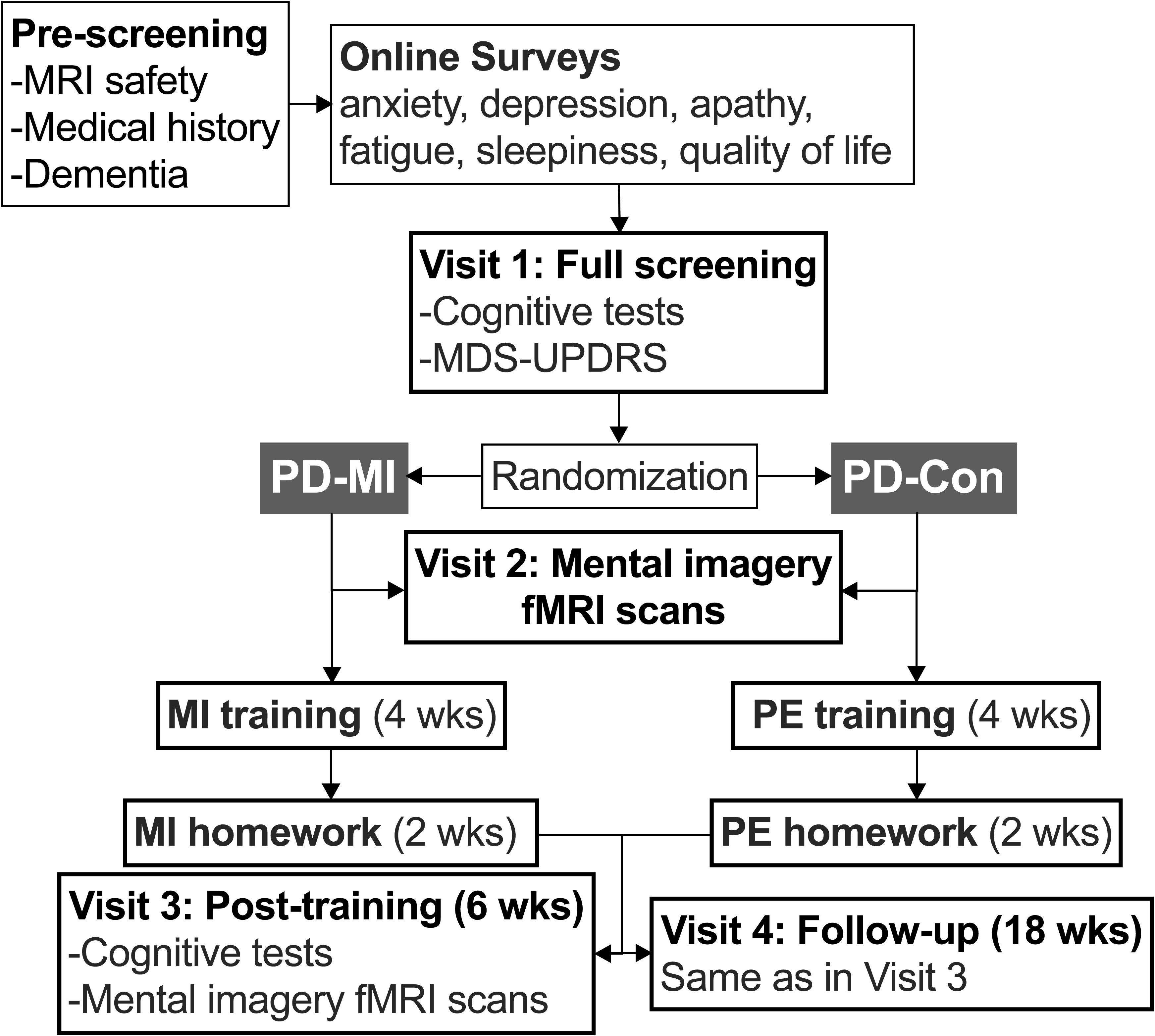
Study flow. After pre-screening, participants took online behavioral surveys, underwent detailed cognitive and motor assessments in the first in-person visit to complete the screening, and were randomized. After baseline scans, participants started their respective trainings and returned for repeat cognitive tests and scans at six weeks post-training and 18 weeks follow-up. FMRI: Functional Magnetic Resonance Imaging, MDS-UPDRS: Movement Disorders Society Parkinson’s disease Unified Rating Scale, MI: Mental imagery, PD: Parkinson’s disease PE: Psychoeducation.

### Participants

We recruited participants with PD via the Yale Movement Disorders Clinics, Adams Comprehensive PD and Movement Disorders Care Center, and local PD support groups between January 2023-July 2024. All participants gave written informed consent in accordance with the procedures approved by the Human Research Protection Office of the Yale School of Medicine.

### Eligibility criteria

People with a diagnosis of PD according to the Movement Disorders Society (MDS) diagnostic criteria^35^ who were 40 years of age or older (idiopathic PD is rare below this age^36^) were included. Metallic implant in the body, claustrophobia, pregnancy, central nervous system disorders other than PD, psychiatric disorders (other than comorbid anxiety and depression), focal findings on neurological exam, Montreal Cognitive Assessment (MoCA)^37,38^ test score < 21/30 (threshold for dementia),^39^ PD-Cognitive Functional Rating Scale (PD-CFRS) score > 4 (cognitive functional ability threshold for dementia),^40^ Hoehn & Yahr (H&Y) disease stage > 3 (stage 3 corresponds to bilateral disease with some imbalance, but physically independent),^41^ an index score that was ≥ 2 standard deviations below the norm on at two or more cognitive tests according to the MDS Level II criteria (represented by either two impaired tests in one cognitive domain or one impaired test in two different cognitive domains)^42^ were the exclusion criteria. The only exception that was made after trial commencement was the delayed memory index score (see below), for which the exclusory cutoff was 1.5 standard deviations below the norm. We chose this stricter threshold to ensure that participants did not have significant memory impairment that could affect their MI performance.^43^

### Sample size calculation

We determined the total sample size of 30 participants (15 in each group) in this pilot trial based on pragmatic considerations regarding recruitment within the set timeframe and budget constraints.^44^ Here, we provide an example of a sample size calculation for a fully powered trial using the Neuro-QoL-CF survey scores as the primary cognitive outcome: The minimal important change in patient-reported outcome measures such as the Neuro-QoL-CF was estimated as 2-6 T-score points.^45^ For example, assuming a change of 4 T-score points on the Neuro-QoL-CF, using the repeated-measures ANOVA within-between factor interaction model with two groups and two measurements, effect size f=0.16 (corresponding to a small-to-moderate effect size d=0.4), alpha 0.05, power 0.80 in G*Power 3.1,^46,47^ a total sample size of 80 (40 in each group) would be needed.

### Screening

Participants underwent screening for MRI safety and medical history. We video-administered the MoCA and PD-CFRS. Participants who passed the initial screening completed online surveys via the REDCap platform to evaluate the contribution of nonmotor factors to cognitive functioning. These included The Geriatric Depression Scale,^48^ Parkinson Anxiety Scale,^49^ Starkstein Apathy Scale,^50^ PD Quality of Life Questionnaire-39,^51^ Parkinson Fatigue Scale,^52^ and the Scales for Outcomes in PD – Sleep.^53^

Participants came for their first in-person visit to Yale to complete the screening. A movement disorders neurologist (S.T.) performed the neurological and movement examination using the MDS-Unified PD Rating Scale (MDS-UPDRS),^54^ which included H&Y staging. Trained research assistants (J.C., A.N., and L.R.) administered cognitive screening tests including the Wechsler Test of Adult Reading (WTAR)^55^ to determine premorbid IQ and the Repeatable Battery for the Assessment of Neuropsychological Status (RBANS)^56^-form B to evaluate attention, delayed verbal/visual memory, language, and visuospatial function; and tests of executive function including the Stroop test,^57^ letter fluency (F-A-S),^58^ and Trail Making Test parts A and B.^59^ The index scores of each RBANS domain and the T-scores of the executive function tests were scored immediately to determine eligibility. Eligible participants completed the computer adaptive version of Neuro-QoL-CF to assess self-reported cognitive functioning in everyday life,^34^ and the Questionnaire on Mental Imagery (QMI) to assess MI skills.^60^

### Randomization and masking

We randomized the eligible participants using the “minimization” method with a 1:1 allocation ratio.^61^ After randomly allocating the first ten participants, each subsequent participant was allocated to either group considering the factors age and sex. This method allowed us to minimize the imbalance between the groups in terms of demographic characteristics. The team member who conducted the initial cognitive assessments was masked to the group assignment of the participants (Figure 2).

**Figure 2.**
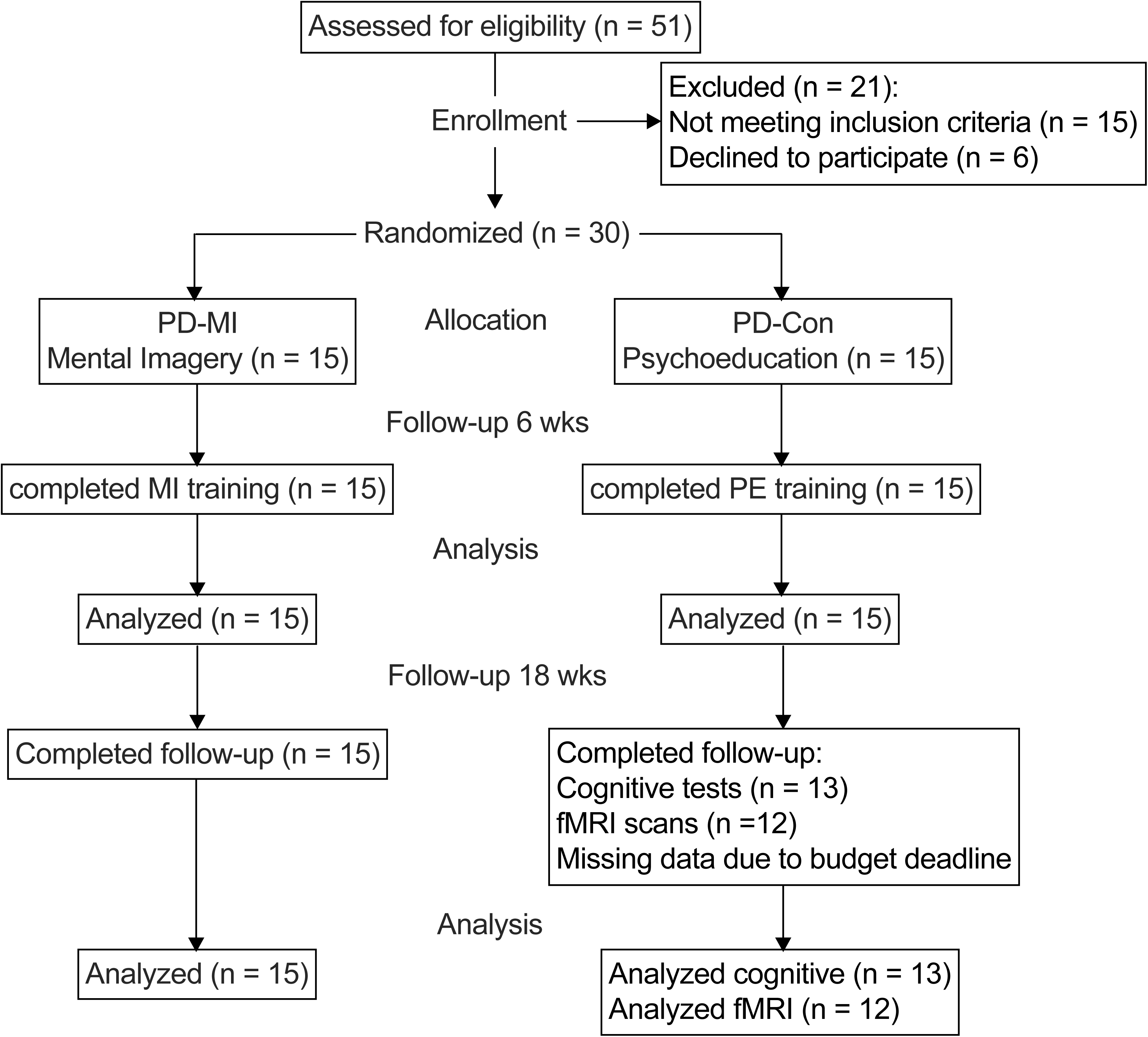
The CONSORT diagram.

### Mental imagery script generation for fMRI scans

We created four MI categories of everyday life: (1) household tasks (e.g., preparing a meal, organizing a closet), (2) outdoor activity (e.g., trip to the grocery store, post office, yard work), (3) event planning (e.g., planning a vacation, party, charity event), and (4) social engagement (e.g., celebration with family, dinner with friends). Participants found a task from each category that was personally relevant and described the main steps of the task to create purposeful, realistic, and vivid scripts. These steps served as the building blocks of the script. Next, we asked participants to elaborate on the steps and elicited episodic details (e.g., when, where, what, who), sensory details (e.g., visual, auditory, tactile, etc.), bodily sensations, actions, feelings, and thoughts associated with the task. Once the final script was created and agreed upon, participants practiced imagining the scripts with closed eyes before going in the scanner. This process was repeated for all four tasks. Participants imagined the same scripts at subsequent visits.

### Scanning

We conducted the fMRI experiments in a Siemens Prisma 3.0 Tesla magnetom with a 64-channel head coil. First, T1-weighted high-resolution MPRAGE anatomical images (voxel size: 1 x 1 x 1 mm) were collected for an accurate localization of the fMRI data. Then, axial, T2*-weighted, echo planar functional images (129 volumes) were collected (voxel size: 3.5 x 3.5 x 4 mm, 36 slices, FoV: 224 mm, TR: 2000 ms, TE: 25 ms, flip angle: 90°). Participants read the task instructions and questions that were displayed on the projector screen through a mirror attached to the head coil. There were four MI runs each lasting 4 min and 18 s. Each run started (for 30 s) and ended (for 40 s) with the baseline task of counting backwards from a randomly selected number while fixating at a cross on the screen. This low-level baseline task was chosen to minimize the possibility of engaging in mind-wandering. After the first baseline task, participants were instructed (6 s) which MI task they were supposed to perform for 3 min with eyes closed. Participants received an auditory signal (2 s) to open their eyes after 3 min. Following each scan, participants answered questions to rate their imagery performance on a scale from 1 to 4 using a button box. These questions included: (1) How vivid was your imagery? (2) How difficult was it to imagine the task? (3) Were you able to plan and implement the task step-by-step in your mind’s eye? (4) Were you able to imagine the setting, people, sensations, emotions, and movements?

After the baseline fMRI experiment, participants started their respective training.

### Mental imagery training

We used the same four categories evenly (event planning, household tasks, outdoor chores, and social engagement) during one-on-one training via Zoom three times a week for the first four weeks (total of 12 sessions). The training sessions were scheduled at participant’s convenience and lasted on average 20 min except for the first session that lasted approximately 45 min. Participant sat in a chair comfortably in a quiet room facing the Zoom camera. We developed the imagery scripts as explained in the previous section. We induced episodic specificity and encouraged interweaving of multisensory details, bodily sensations, actions, thoughts, and feelings associated with the task. The scripts were further enriched by incorporating event and time markers as prospective memory cues (e.g., remember to stop by the post office on the way to the grocery store), potential obstacles and ways to overcome them (e.g., road closure and detour on the way to the store), and self-monitoring strategies (e.g., check that all items on the list have been purchased). Upon completion of the script, we guided participant through breathing (deep inhalation/exhalation, four times) and stretching exercises (neck, shoulders, arms) and instructed participant to start imagining the script with eyes closed at their own pace. We started the timer, turned our Zoom camera off and muted the microphone to avoid distraction. When participant was done, researcher wrote down the duration of practice. Subsequently, participant rated their imagery experience on a scale from 1 to 4 as explained in the previous section. Participants also practiced MI on their own twice a week (on the no-training days) in the first four weeks and five days a week in the subsequent two-week block. After each of these self-practices, participants filled out a detailed diary via the REDCap online platform. We structured this diary to capture the key elements of MI including the setting and components of the activity; contextual, sensory, and episodic details; thoughts and emotional experiences, and vividness and difficulty level of imagery.

### Psychoeducation

Participants in the PD-Con group received psychoeducation three times a week for four weeks on cognitive functioning and brain health in PD. We recorded 15-min lecture modules during which participants watched a slide presentation and listened to the instructor’s voice. After the lecture, participants completed short quizzes for approximately 5 min. The modules covered topics including brain anatomy, neuroplasticity, attention-executive function, memory, speech-communication, hallucinations and delusions, mood disorders, stress, and social bonds, as well as the role of sleep, diet, physical activity, and mind-body interventions in managing the cognitive and mood symptoms. On two no-lecture days each week in the first four weeks, then daily for five days a week in the subsequent two-week block participants completed quizzes about lecture contents via the REDCap platform.

### Post-training visits

Upon completion of the six-week training, all participants returned for repeat cognitive evaluations and fMRI scans. After this visit, we encouraged participants in the PD-MI group to incorporate goal-directed MI into their daily life and fill out a log to indicate their practice frequency until the final visit at 18 weeks. We encouraged participants in the PD-Con group to retain the psychoeducational information and apply the recommendations about brain and cognitive health.

At 18 weeks, participants returned to repeat the cognitive evaluations and fMRI scans.

### Subjective reports of cognitive change

We created a semi-structured survey to assess the subjective changes in multiple cognitive domains including attention, processing speed, executive function, memory, visuospatial skills, language, and social cognition, and applied it to a subset of participants from both groups at six weeks. Participants rated the subjective changes as none (0), slight (1), mild (2), moderate (3), or significant (4) (see Supplement 2).

### Experience with MI practice

At the end of weeks five and six, then biweekly during the 12-week self-practice periods, we conducted brief phone interviews with participants in the PD-MI group to assess their experiences with the practice.

## Data Analysis

### Analysis of cognitive and imagery data

We obtained a composite executive function score by averaging the T-scores on the Stroop, letter fluency, and Trail Making-B tests. We calculated the WTAR scores and total and domain-specific RBANS index scores. The Neuro-QoL-CF score was calculated automatically by the computer program. We averaged the MI performance ratings during the training period for the PD-MI group and during fMRI scans for both groups and created a MI vividness score.

### Preprocessing of fMRI data

We used the CONN toolbox for all fMRI data analysis steps.^62^ We performed motion correction, outlier detection, coregistration of functional scans with the anatomical scan, segmentation and normalization to the standard MNI template, and smoothing with an 8-mm kernel to account for inter-individual anatomical variability. De-noising steps included the elimination of signal originating from the white matter and cerebrospinal fluid using the CompCor method,^63^ regression of motion artifacts and outliers (i.e., frame-wise displacement above 1.5 mm or global signal changes above 7 standard deviations) from the time series, linear detrending, and high-pass filtering (0.008 Hz < f < Inf).

### Functional connectivity analysis

To compare the task-specific functional connectivity changes between the groups, we used the generalized psychophysiological interaction (gPPI) model. We first convolved the 3-min task blocks in each MI run with the canonical hemodynamic response function. We chose the functionally defined main regions of interest (ROIs) (N = 32) of the major networks in the CONN toolbox including the default mode, frontoparietal, dorsal attention, salience, language, sensorimotor, visual, and cerebellar networks. For each participant, we extracted the average blood oxygenation level-dependent signal time courses calculated via the gPPI model from these nodes and correlated them with each other using Pearson correlations. The r values corresponded to the functional connectivity strength between node pairs. We Fisher z-transformed the r values to obtain group-level functional connectivity maps for statistical analyses.

### Statistical Analyses

#### Outcome measures

For our primary objective, the primary and secondary behavioral outcome measures were change in the Neuro-QoL-CF and composite executive test scores at six weeks, respectively. Change in the RBANS subdomain index scores at six weeks were the exploratory behavioral outcome scores. Change in network functional connectivity during the MI tasks at six weeks was the primary imaging outcome. For our secondary objective, we used the same behavioral and imaging outcome measures at 18 weeks. We also used the change in MI vividness scores in the scanner at six and 18 weeks as manipulation check.

#### Demographic, clinical, and nonmotor symptom data

We performed independent-sample t-tests to compare the normally distributed continuous baseline data and nonparametric Mann-Whitney U tests to compare the discrete baseline data and non-normally distributed continuous data between the PD-MI and PD-Con groups (p < 0.05, two-tailed) using the SPSS Version 29. The disease duration and WTAR scores were significantly different between the groups (see Results) and were included as covariates in the repeated measures ANCOVA tests.

#### Cognitive and imagery data

For our primary objective, we compared the cognitive data of the two groups at baseline and six weeks using a repeated measures ANCOVA (dependent variables: cognitive test scores, MI quality scores; between-subject factor: group, within-subject factor: time with two levels, interaction: group-by-time). For our secondary objective, we used the same ANCOVA approach with time as the within-subject factor with three levels.

We also explored the relationship between the changes in the Neuro-QoL-CF and MI vividness scores using simple correlations.

#### fMRI data

For our primary objective, we compared the pairwise network connectivity during MI tasks between the two groups at baseline and six weeks using a repeated measures ANOVA (dependent variables: connectivity measures, between-participant factor: group, within-participant factor: time with two levels, interaction: group-by-time) using the CONN toolbox. For our secondary objective, we used the same ANOVA approach with time as the within-subject factor with three levels. We used FDR-correction for multiple comparisons (p < 0.05) followed by post hoc tests.

## Results

### Baseline demographic, clinical, and nonmotor data

All participants had mild bilateral disease (H&Y disease stage 2). The PD-MI group had significantly higher disease duration and higher WTAR scores compared to the PD-Con group. The PD-Con group had higher education that reached a trend toward statistical significance. The groups were well matched in all other baseline characteristics (Table 1 and Table S1 in Supplement 2).

**Table 1.** Demographic and baseline clinical data.

|  | PD-MI<br>(N=15) | PD-Con<br>(N=15) | P value |
| --- | --- | --- | --- |
| Age | 67.6 ± 8.6 | 67.5 ± 7.7 | 0.956 |
| Sex | 4 F, 11 M | 3 F, 12 M | 0.679 |
| Education (yrs) | 17.1 ± 1.9 | 18.3 ± 1.8 | 0.066 |
| Disease duration (yrs) | 7.0 ± 3.1 | 3.7 ± 3.1 | 0.010* |
| MDS-UPDRS I | 8.1 ± 5.6 | 8.4 ± 5.9 | 0.876 |
| MDS-UPDRS II | 6.1 ± 4.7 | 6.4 ± 4.2 | 0.713 |
| MDS-UPDRS III | 25.2 ± 8.2 | 26.1 ± 8.0 | 0.754 |
| MDS-UPDRS IV | 0.9 ± 2.0 | 1.6 ± 3.0 | 0.412 |
| MDS-UPDRS total | 40.3 ± 10.8 | 42.5 ± 13.9 | 0.621 |
| MoCA | 27.8 ± 1.6 | 27.4 ± 1.5 | 0.475 |
| PD-CFRS | 1.4 ± 1.3 | 1.7 ± 1.4 | 0.512 |
| GDS | 5.6 ± 5.9 | 5.1 ± 4.8 | 0.967 |
| PAS total | 7.7 ± 7.5 | 8.7 ± 6.7 | 0.713 |
| Apathy | 11 ± 7.1 | 11.8 ± 5.3 | 0.729 |
| PFS-16 | 2.2 ± 0.9 | 2.5 ± 1.1 | 0.354 |
| SCOPA-sleep | 9.7 ± 5.7 | 9.8 ± 7.9 | 0.653 |
| PDQ-39-SI | 13.1 ± 9.4 | 14.9 ± 9.5 | 0.486 |
| WTAR | 118.2 ± 9.4 | 116.3 ± 4.7 | 0.041* |
| QMI | 70.3 ± 24.9 | 64.7 ± 19.3 | 0.497 |
| LEDD (mg) | 378.7 ± 324.2 | 488.9 ± 318.7 | 0.389 |

\*nonparametric Mann-Whitney U test
GDS: Geriatric Depression Scale, LEDD: Levodopa equivalent daily dose,<sup>64</sup> MDS-UPDRS: Movement Disorders Society Unified Parkinson's Disease Rating Scale (Higher scores indicate more disability. Part I: Nonmotor aspects of experiences of daily living, part II: Motor aspects of experiences of daily living, part III: Motor examination, part IV: Motor complications). MoCA: Montreal Cognitive Assessment test, PAS: Parkinson Anxiety Scale, PD-CFRS: Parkinson's Disease Cognitive Functional Rating Scale. PDQ-39-SI: Parkinson's disease questionnaire (quality of life) summary index, PFS-16: Parkinson's Disease Fatigue Scale (average), QMI: Questionnaire on Mental Imagery, SCOPA-sleep: Scales for Outcomes in Parkinson's Disease – Sleep, WTAR: Wechsler Test of Adult Reading.

### Training

Both groups were fully compliant with the six-week training requirements. During the subsequent 12-week period, the PD-MI group practiced MI on average three days a week (Table S2 in Supplement 2).

### Post-training cognitive changes

The scores are listed in Table 2. There was no significant two-way or three-way interaction between WTAR, disease duration, and group. At six weeks, the full factorial model with WTAR and disease duration as covariates showed a significant group-by-time interaction in Neuro-QoL-CF scores (F(1,26) = 6.802, p = 0.015, partial 11^2^ = 0.207) (Figure 3A). There was a significant group-by-time interaction in the average MI vividness ratings in the scanner (F(1,26) = 5.000, p = 0.034, partial 11^2^ = 0.161) with higher ratings in the PD-MI group post-training (Figure 3B). There was no significant group-by-time interaction in composite executive function scores (F(1,26) = 0.431, p = 0.517) or in RBANS subdomain scores (Table S3 in Supplement 2). At 18 weeks, there were no significant group-by-time interactions in any of the outcome measures (Table S3 in Supplement 2). As an exploratory analysis, we also compared the average performance ratings after the first and fourth weeks of the MI training in the PD-MI group using a paired-sample t-test and found a trend toward significance (p = 0.056).

**Figure 3.**
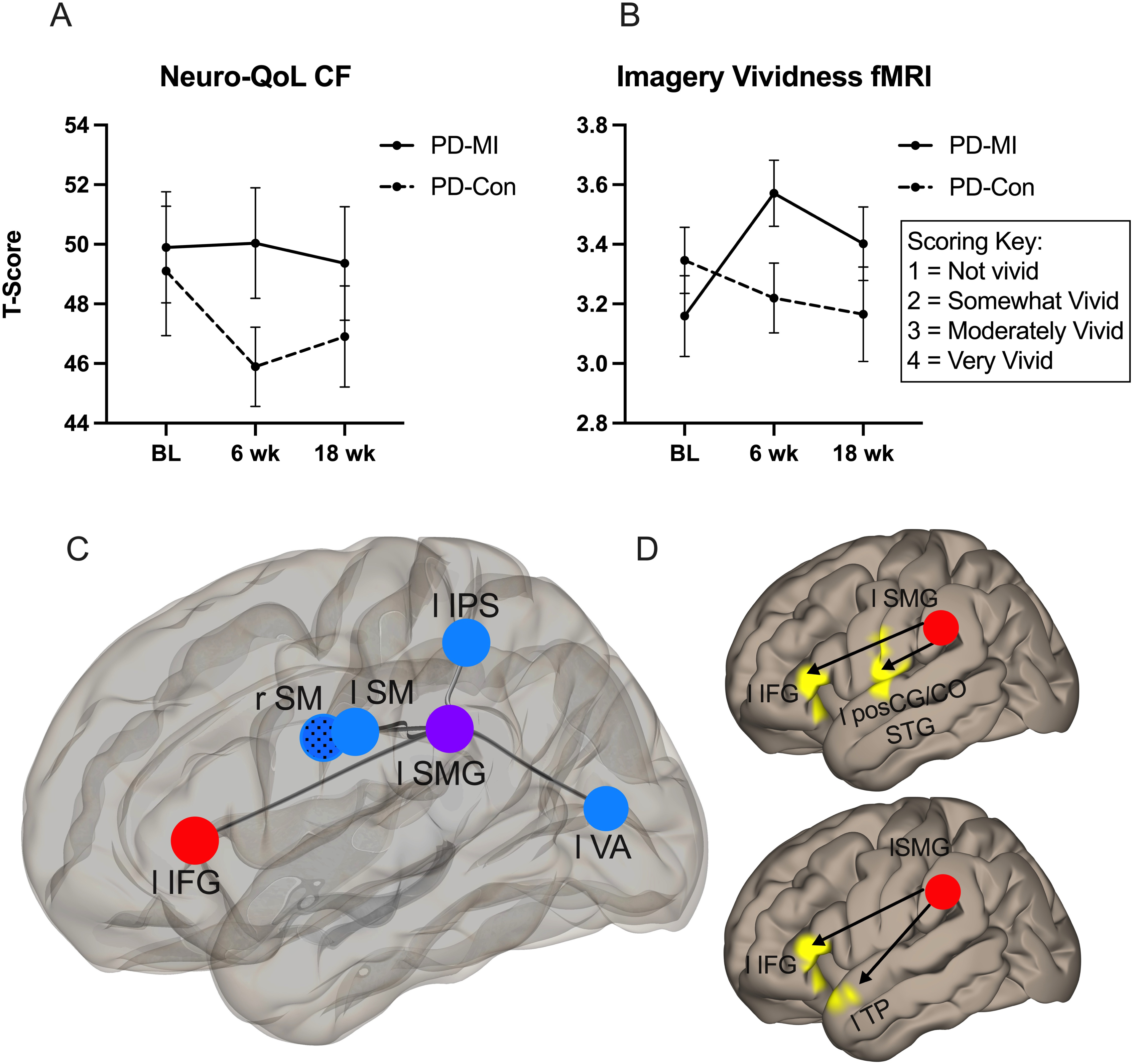
Main outcomes. **Cognitive**: Changes in Neuro-QoL-CF scores (**A**) and imagery vividness ratings during MI tasks in the scanner (**B**). Error bars show standard error. BL: Baseline. **Imaging**: Pairwise functional connectivity differences during MI tasks in the PD-MI > PD-Con contrast at six (red) and 18 weeks (blue) compared to baseline (**C**). Whole-brain voxel-wise functional connectivity differences during MI tasks using the left SMG as the seed in the PD-MI > PD-Con contrast at six and 18 weeks (**D**). *Top*: F-test results (any difference), *bottom*: Post hoc six-week > baseline comparison. CO: Central operculum, IFG: Inferior frontal gyrus, IPS: Intraparietal sulcus, postCG: Postcentral gyrus, SM: Sensorimotor, SMG: Supramarginal gyrus, STG: Superior temporal gyrus, TP: Temporal pole, VA: Visual association cortex.

**Table 2.** Cognitive data.

|  | **Baseline** |  | **6-wk** |  | **18-wk** |  |
| --- | --- | --- | --- | --- | --- | --- |
|  | PD-MI | PD-Con | PD-MI | PD-Con | PD-MI | PD-Con<sup>^</sup> |
| Neuro-QoL-CF | 49.9 ± 7.2<br>(39.5 – 64.1) | 49.1 ± 8.4<br>(40.9 – 68.3) | 50.0 ± 7.2<br>(42.6 – 64.5) | 45.9 ± 5.1<br>(38.3 – 55.9) | 49.4 ± 7.4<br>(40.6 – 68.3) | 46.9 ± 6.1<br>(38.1 – 56.2) |
| *RBANS:* |  |  |  |  |  |  |
| Visuospatial | 103.5 ± 14.2 (72 – 131) | 97.1 ± 12.1 (66 – 121) | 100.8 ± 16.5 (72 – 131) | 96.3 ± 17.7 (64 – 131) | 101.9 ± 18.6 (62 – 126) | 96.4 ± 13.3 (72 – 112) |
| Language | 99.6 ± 9.2 (85 – 117) | 99.3 ± 10.7 (87 – 131) | 101.9 ± 12.4 (87 – 131) | 99.5 ± 6.9 (87 – 112) | 101.7 ± 12.2 (85 – 131) | 100.2 ± 11.0 (82 – 128) |
| Attention | 108.0 ± 15.6 (72 – 128) | 107.3 ± 10.4 (88 – 122) | 111.3 ± 13.9 (85 – 132) | 107.9 ± 12.5 (91 – 125) | 112.3 ± 14.4 (91 – 135) | 107.3 ± 13.3 (85 – 125) |
| Memory | 100.3 ± 11.6 (81 – 121) | 99.8 ± 10.5 (84 – 116) | 105.1 ± 14.2 (71 – 126) | 106.9 ± 13.1 (81 – 126) | 112.1 ± 10.3 (95 – 126) | 107.9 ± 9.9 (88 – 122) |
| *Executive tests:* |  |  |  |  |  |  |
| Composite | 53.1 ± 7.9 (38.0 – 71.3) | 48.7 ± 7.4 (36.3 – 66.7) | 54.7 ± 8.2 (40.3 – 73.0) | 51.0 ± 9.5 (30.3 – 69.3) | 55.1 ± 9.6 (35.7 – 72.7) | 49.5 ± 7.8 (36.7 – 63.7) |
| TMT-B | 50.9 ± 12.0 (33 – 81) | 47.5 ± 11.9 (31 – 80) | 57.0 ± 11.9 (37 – 81) | 49.8 ± 13.7 (27 – 84) | 55.1 ± 13.2 (33 – 72) | 47.5 ± 11.0 (27 – 62) |
| Letter fluency (F-A-S) | 53.7 ± 10.3 (30 – 70) | 47.5 ± 11.1 (28 – 65) | 54.5 ± 10.9 (30 – 73) | 51.0 ± 11.7 (35 – 72) | 55.2 ± 12.7 (23 – 73) | 48.9 ± 11.5 (35 – 72) |
| Stroop | 54.6 ± 8.8 (44 – 78) | 51.0 ± 6.9 (41 – 62) | 52.7 ± 6.1 (42 – 65) | 52.3 ± 8.6 (27 – 64) | 55.0 ± 8.7 (42 – 80) | 52.2 ± 7.6 (42 – 69) |

The means ± standard deviations (range) of T values for Neuro-QoL-CF and executive function tests and of index scores of RBANS domains are listed.
<sup>^</sup>At 18 weeks, data of 13 participants in the PD-Con group were available.
Neuro-QoL-CF: Quality of Life in Neurological Disorders Cognitive Function survey (version 2), RBANS: Repeatable Battery for the Assessment of Neuropsychological Status, TMT-B: Trail Making Test – B.

The exploratory correlation analyses between the change in MI vividness score during fMRI scans and in Neuro-QoL-CF scores at six weeks showed a significant positive correlation in the PD-MI group (Pearson r = 0.525, p =0.044), but not in the PD-Con group (non-normal distribution, Spearman rho = 0.036, p = 0.899). The correlation between the change in MI vividness scores at four weeks of training and in Neuro-QoL-CF scores at six weeks in the PD-MI group was also significantly positive (Pearson r = 0.609, p =0.016).

### Subjective reports of cognitive change

Ten participants in the PD-MI and eight participants in the PD-Con group completed the surveys at six weeks. In the PD-MI compared to the PD-Con group, there was a slight but significant improvement in the executive functioning, memory, social cognition, and all cognitive domains averaged (see Table S7 in Supplement 2).

### Experience with MI practice

Briefly, improvement in planning and organizing daily tasks, increased awareness of surroundings, and better self-regulation were most reported positive changes. These changes were consistent with those reported in the subjective cognitive function survey. Finding quiet time and different themes to practice were reported as challenges (Table S4 in Supplement 2).

### Imaging results

There was minimal head motion during scans which did not differ significantly between the groups (Table S5 in Supplement 2). The functional connectivity results are summarized in Table 3. The details of the MI scenarios practiced in the scanner are summarized in Table S6 in Supplement 2.

**Table 3.** Functional connectivity results.

| **ROI-to-ROI 6-wk changes** | ROI 1 | ROI 2 | T (28) | p-FDR |
| --- | --- | --- | --- | --- |
| *Main effect of group:* |  |  |  |  |
| PD-MI, MI2 > MI1 | R AG | MPFC | 4.1 | 0.009 |
| PD-Con, MI2 > MI1 | L SMG | R SM | 3.6 | 0.040 |
| *Group-by-time interaction:* |  |  |  |  |
| PD-MI vs PD-Con, MI2 > MI1 | L IFG | L SMG | 4.1 | 0.009 |
| **ROI-to-ROI 18-wk changes** | ROI 1 | ROI 2 | F (2,27) | p-FDR |
| *PD-MI vs PD-Con, any difference between MI1, MI2, and MI3* | L SMG | L SM | 12.8 | 0.004 |
|  | L SMG | L VA | 8.8 | 0.014 |
|  | L SMG | R SM | 8.6 | 0.014 |
|  | L SMG | L IPS | 7.4 | 0.021 |
|  | L IFG | L SMG | 8.5 | 0.044 |
| *Post hoc comparisons:* | ROI 1 | ROI 2 | T (28) | p-FDR |
| PD-MI vs PD-Con, MI2 > MI1 | L IFG | L SMG | 4.1 | 0.009 |
| PD-MI vs PD-Con, MI3 > MI1 | L SMG | L SM | -4.9 | 0.001 |
|  | L SMG | R SM | -4.1 | 0.005 |
|  | L SMG | L IPS | -3.9 | 0.006 |
|  | L SMG | L VA | -3.8 | 0.006 |
| PD-MI vs PD-Con, MI3 > MI2 | - | - | - | - |
| **L SMG ROI-to-Voxel** | Regions | Cluster Size | MNI coordinates (x, y, z) | p-FDR |
| *PD-MI vs PD-Con, any difference between MI1, MI2, and MI3 ($F(2,50)_{min} = 5.06$)* | L vLPFC | 440 | -56, 32, 6 | 0.007 |
|  | L postCG, CO, STG | 415 | -58, -8, 2 | 0.007 |
| *Post hoc comparisons $T(25)_{min} = 2.79$:* |  |  |  |  |
| PD-MI vs PD-Con, MI2 > MI1 | L vLPFC | 631 | -48, 26, -4 | 0.001 |
| PD-MI vs PD-Con, MI3 > MI1 | - | - | - | - |
| PD-MI vs PD-Con, MI3 > MI2 | - | - | - | - |

Group comparisons of functional connectivity during MI scans at baseline (MI1), six weeks (MI2) and 18 weeks (MI3). The network regions of interest (ROI) in the CONN toolbox and clusters with their coordinates on the standard Montreal Neurological Institute (MNI) brain template are listed. AG: Angular gyrus (executive network), CO: Central operculum, IFG: Inferior frontal gyrus (language network), IPS: Intraparietal sulcus (dorsal attention network), MPFC: Medial prefrontal cortex (default mode network), postCG: Postcentral gyrus, SM: Sensorimotor cortex (sensorimotor network), SMG: Supramarginal gyrus (salience network), STG: Superior temporal gyrus, TP: Temporal pole, VA: Visual association cortex (visual network), vLPFC: Ventrolateral prefrontal cortex (including the inferior frontal gyrus, orbitofrontal cortex, and frontal operculum).

At six weeks, the main effect of group was increased functional connectivity between the parietal (executive network) and medial prefrontal (default mode network) regions for the PD-MI and between motor and visuospatial regions for the PD-Con group. The functional connectivity between the left inferior frontal gyrus (IFG) and left supramarginal gyrus (SMG) was stronger after six weeks of training in the PD-MI vs PD-Con group (Figure 3C). At 18 weeks compared to baseline, the functional connectivity between the left SMG and several motor and visuospatial ROIs was weaker in the PD-MI compared to the PD-Con group (Figure 3C). A post hoc ROI-to-voxel whole brain functional connectivity analysis using the left SMG as the seed ROI showed increased functional connectivity with ventrolateral prefrontal and temporal regions in the PD-MI compared to the PD-Con group at six weeks (Figure 3D). There were no significant group differences at 18 weeks.

## Discussion

In summary, after six weeks of MI training, there was a significant difference in Neuro-QoL-CF scores and MI vividness in the PD-MI compared to the PD-Con group, but this difference was not sustained at 18 weeks. Notably, changes in MI vividness correlated with changes in Neuro-QoL-CF scores at six weeks only in the PD-MI group further supporting a link between improved MI skills and subjective cognitive functioning. In surveys, the PD-MI group reported improvement in planning and organizational skills, task preparedness, self-regulation, and awareness in daily life all of which are related to frontal executive functions, as well as more benefit in memory and social cognition compared to the PD-Con group, whereas the PD-Con group endorsed problems in these cognitive domains more frequently consistent with the decline in their Neuro-QoL-CF scores. However, we did not find a significant post-training difference in the composite executive test scores (or in test scores of other cognitive domains) between the groups at six or 18 weeks. One possibility is the limited ecological validity of neuropsychological tests as it pertains to everyday cognitive skills.^65^ In other words, performance on these standardized tests administered in controlled experimental settings does not necessarily predict real-world performance. This limitation is particularly important because of the personalized nature of the MI training focusing on everyday cognitive functioning. Therefore, we think that the self-report measures may have provided more valid outcomes for the MI training, which is also an important consideration when selecting cognitive outcome measures in future fully powered trials using similar personalized cognitive interventions. Another possibility is that our PD cohort included community-dwelling, independent adults with mild disease and without significant cognitive impairment. Therefore, standardized cognitive tests may not have been sensitive enough to detect subtle changes in a relatively high-functioning cohort.

The MI training (especially the one-on-one sessions) was well received with the exceptions that finding time was challenging and the homework requirement was disliked by some participants. Compliance with MI practice in the subsequent unsupervised 12-week period was on average three days a week which may have contributed to the loss of benefits during that time. Our results are within the range of durability of outcomes that was reported by physical therapy interventions with similar schedules and underline the challenges with long-term treatment adherence.^66^

Taken together, these findings suggest that consistent daily MI practice may be an effective tool to promote cognitive preparedness for everyday tasks in people with PD. However, potential longer-term sustained benefits and effect of practice frequency of MI need further investigation.

Our imaging results showed stronger functional connectivity between the default mode and executive network regions in the PD-MI group at six weeks consistent with previous reports of mental simulations of goals and plans.^27–31^ On the other hand, PD-Con group showed stronger functional connectivity between motor and visuospatial network regions. There was also a significant increase in the left SMG-IFG functional connectivity in the PD-MI compared to the PD-Con group. The post hoc ROI-to-whole brain voxel-wise connectivity analysis corroborated this increase. Yet, the left SMG showed decreased functional connectivity with motor and visuospatial regions in the PD-MI compared to the PD-Con group at 18 weeks compared to baseline. The MI task involves lower-level visuospatial and motor processes and engages the corresponding networks as we have previously demonstrated.^32,33^ Moreover, the SMG and IFG, which are parts of the ventral visual attention network,^67,68^ are also major hubs that show consistent activation during various motor^69^ and visual imagery tasks.^70^ It is worth noting that the network membership of brain regions is not fixed but varies according to task demands. The SMG is also considered a member of the salience network involved in orienting to salient external stimuli and internal events,^71^ and the left IFG is part of the language network that interacts with several cognitive control networks (e.g., theory of mind, default mode) during language processing.^72^

Our results suggest that as participants master the lower-level sensorimotor processes of MI with practice, a shift to higher-level internal thought processes takes place. The functional reorganization of networks from decreased connectivity in the sensorimotor to increased connectivity in the cognitive networks may reflect the neural basis of this cognitive remapping. Of note, because of the small sample size of this pilot study, we chose only the main nodes of functional networks for the connectivity analyses. A fully powered trial should include network nodes that cover the whole brain. We would also suggest that such a trial should consider multivariate mediation analyses to examine the relationship between network functional connectivity changes and cognitive changes as a result of MI training.

In conclusion, our study shows the feasibility of MI training to support everyday cognitive functioning in people with PD. The MI approach we used here is unique in that it promotes a flexible and implicit strategy that is directly linked to the cognitive tasks of everyday living (i.e., MI of the task facilitates preparedness for the actual implementation of the task). The personalized nature of the MI training ensures that each person’s own repertoire of everyday activities is targeted. We envision that regular MI practice could be a tool to preserve or improve resilience against cognitive functional decline and be further developed as part of a comprehensive personalized neurocognitive rehabilitation program to meet the cognitive functional needs of people with PD or of older adults without PD who experience similar everyday cognitive deficits. The strengths of our study are the detailed clinical and cognitive characterization of our well-matched PD cohorts, consistent and practical delivery of the supervised MI training on a teleconference platform showing the feasibility of remote training, and automated processing of the primary cognitive and imaging outcomes in an unbiased way. Our study has several limitations. First, it is a pilot study with a relatively small sample size, therefore, even the statistically significant results should be interpreted as preliminary and not used in sample size calculations. We provided a sample size estimate for a future fully powered trial. Second, we showed short-term subjective benefits of MI. Strategies to improve the long-term effects of MI (e.g., appropriate duration and frequency of training, delivery of training – supervised versus unsupervised, individual or group setting, etc.) need further investigation. Third, our relatively high-functioning PD cohort was capable of practicing MI. The proposed MI protocol may not be suitable for cognitively impaired people with PD, for example, for those with significant memory deficits.^43^ The required modifications of MI training based on the types and severity of cognitive deficits in people with PD need to be addressed in future studies.

## Data Availability

All data produced in the present study are available upon reasonable request to the authors

## Acknowledgments

The authors thank the study participants for their participation and the Yale Movement Disorders neurologists, Drs. Veronica Santini, Amar Patel, Sara Schaefer, and Hae Hawong, for their assistance in participant recruitment.

## Ethical approval

This study (protocol # HIC2000033352) was approved by the Human Research Protection Office of the Yale School of Medicine

---

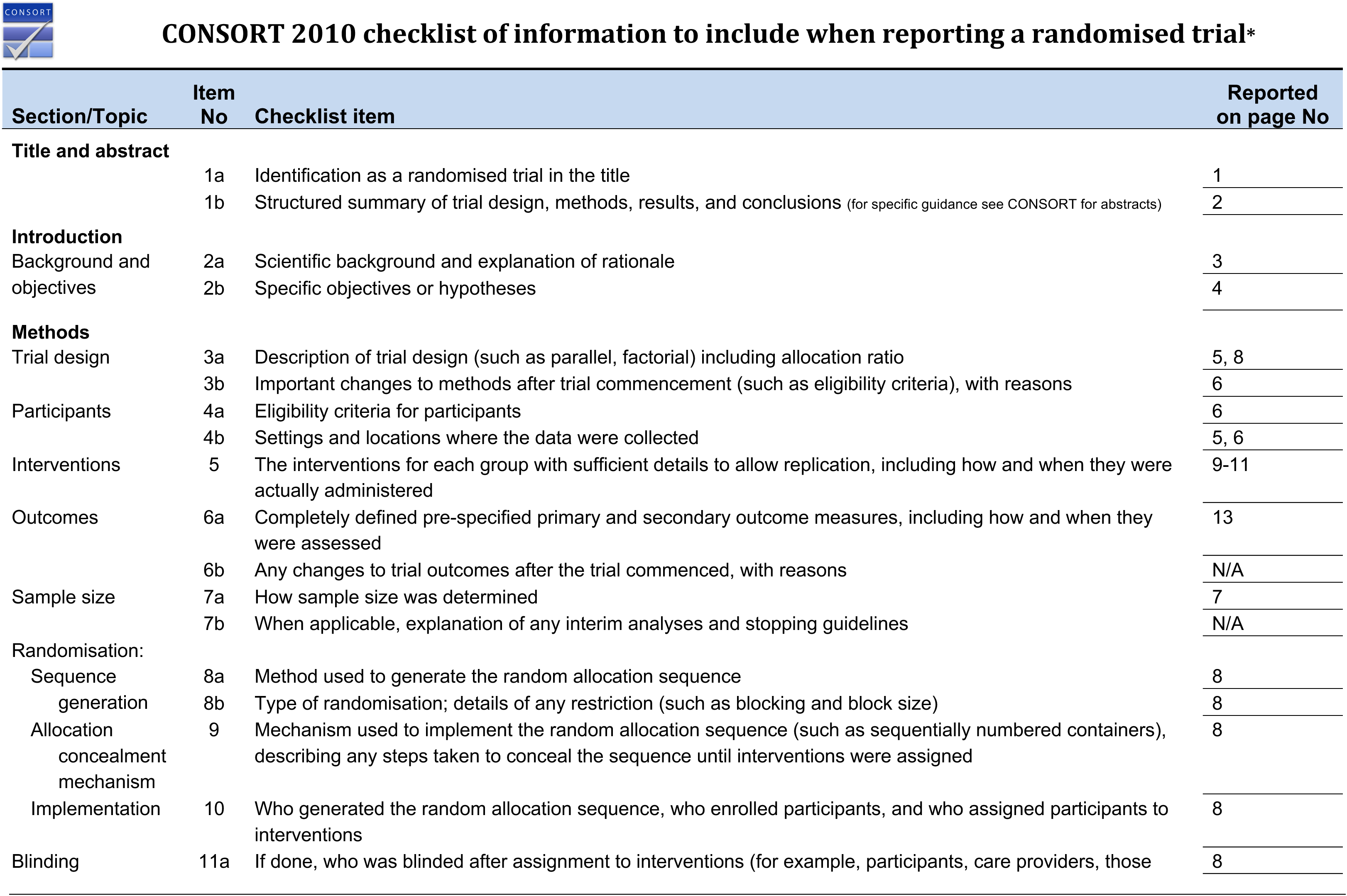
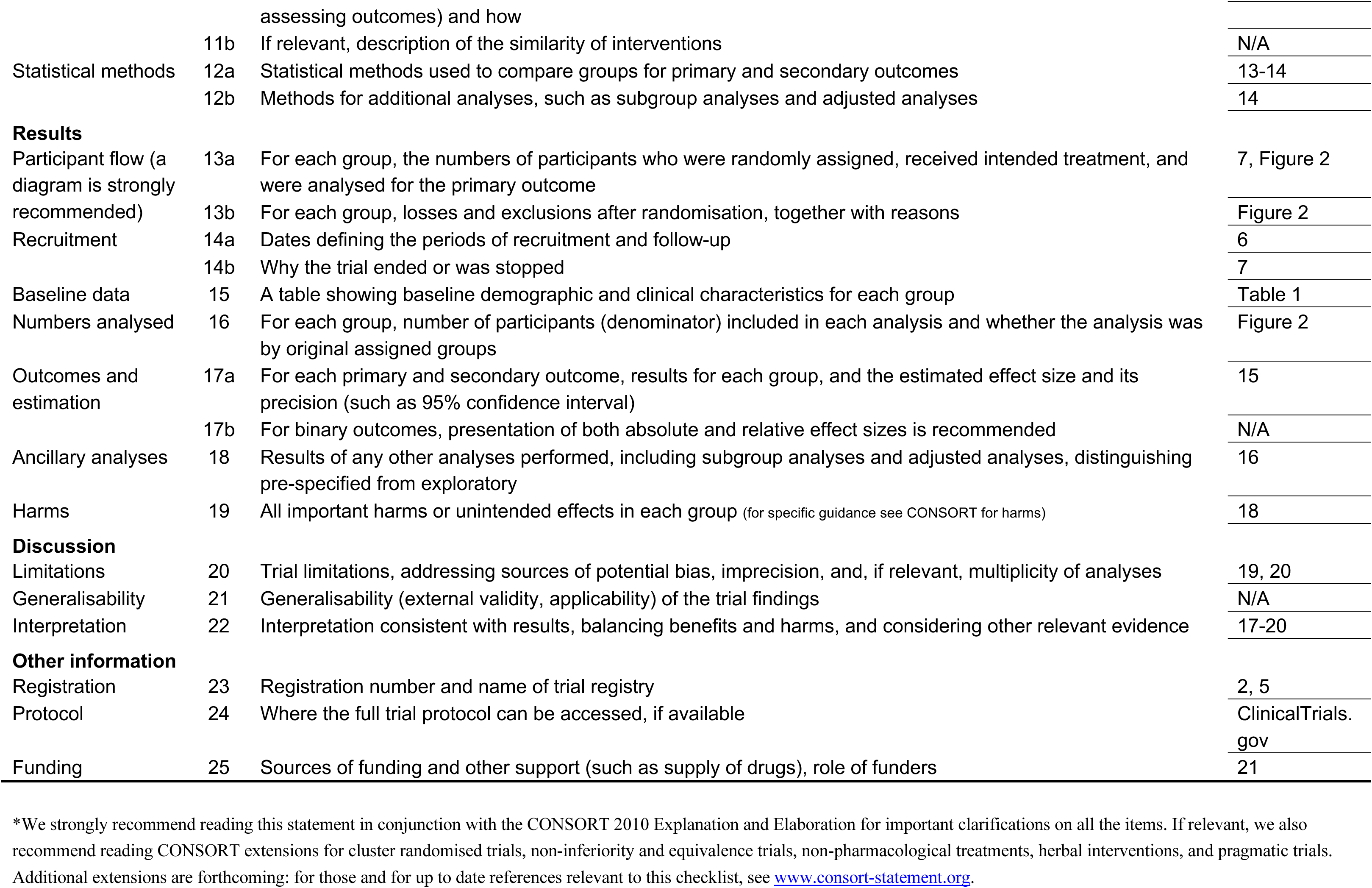

## Supplementary Material 2

We performed one-sample t-tests comparing the means of our PD cohort to the cutoff scores or population means for PD patients. Briefly, our PD cohort showed significantly lower depression, anxiety, and apathy scores. Quality of life and nighttime sleep scores were significantly better.

Daytime sleepiness scores were not significantly different (or were better) depending on the cutoff.

**Table S1.**
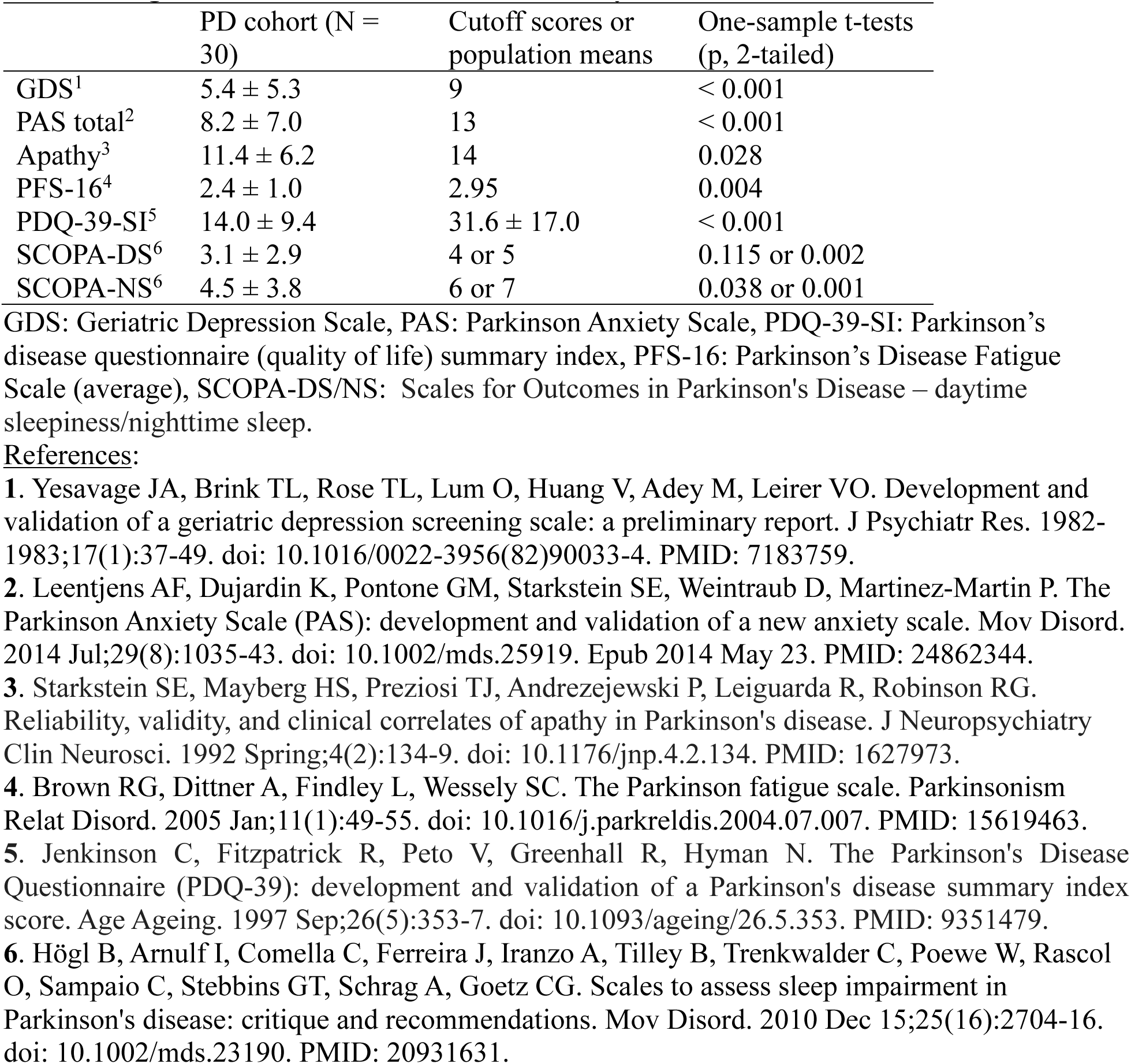
Significance of baseline nonmotor survey scores.

**Table S2.** Practice and homework compliance.

|  | 6-wk period | 12-wk period |
| --- | --- | --- |
| **PD-MI practice frequency** |  |  |
| *Category:* |  |  |
| Event planning | 6.1 | 8.8 |
| Household task | 7.5 | 8.3 |
| Outdoor task | 8.4 | 11.6 |
| Social event | 7.9 | 6.8 |
| Total | 29.9 | 35.5 |
| **PD-Con accuracy in quizzes** | 90% ± 12% | - |

The maximum total number of required MI practices in the 6-week period was 30 (12 one-on-one training sessions and 18 self-practice). The PD-MI group on average completed 29.9 practices (including all 12 one-on-one sessions). The average duration of MI practice during the one-on-one sessions was 5.0 min ± 1.3 min. In the subsequent 12-week period, participants were encouraged to practice MI daily and maintain a log. The PD-MI group practiced MI on average 3 times per week. One subject did not keep a log but reported the same average frequency. The PD-Con group was fully compliant with lecture requirements and completed all quizzes assigned in the 6-week period.

**Table S3.** Nonsignificant post-training changes.

|  | 6 weeks | 18 weeks |
| --- | --- | --- |
|  | time-by-group interaction | time-by-group interaction |
| Composite executive |  | F(2,48) = 0.171, p = 0.844 |
| Neuro-QoL |  | *F(2,48) = 2.529, p = 0.104 |
| MI vividness |  | *F(2,48) = 2.689, p = 0.103 |
| *RBANS index scores:* |  |  |
| Visuospatial | F(1,26) = 0.005, p = 0.943 | F(2,48) = 0.049, p = 0.952 |
| Language | F(1,26) = 0.020, p = 0.888 | F(2,48) = 0.080, p = 0.923 |
| Attention | F(1,26) = 1.370, p = 0.252 | F(2,48) = 2.031, p = 0.142 |
| Memory | F(1,26) = 0.241, p = 0.627 | F(2,48) = 2.071, p = 0.137 |

WTAR and disease duration were used as covariates in both ANCOVA models.
\*Sphericity assumption was not met. The p value is Greenhouse-Geisser corrected.

**Table S4.**
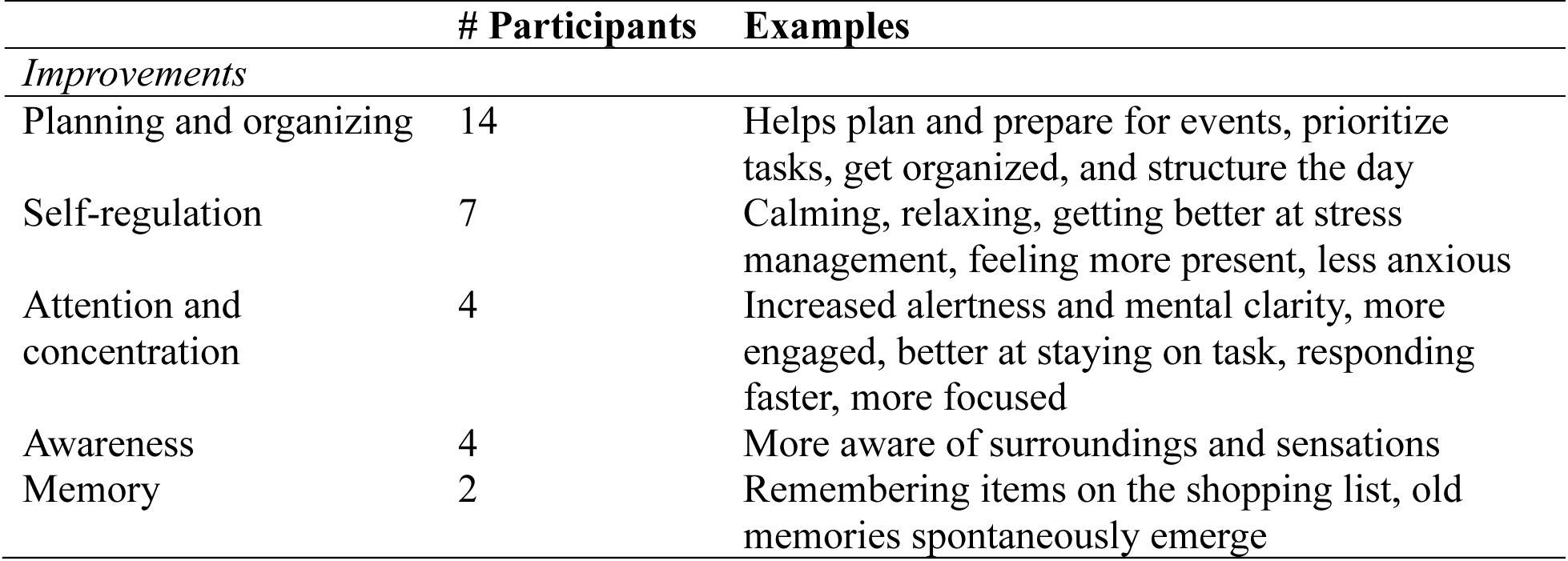
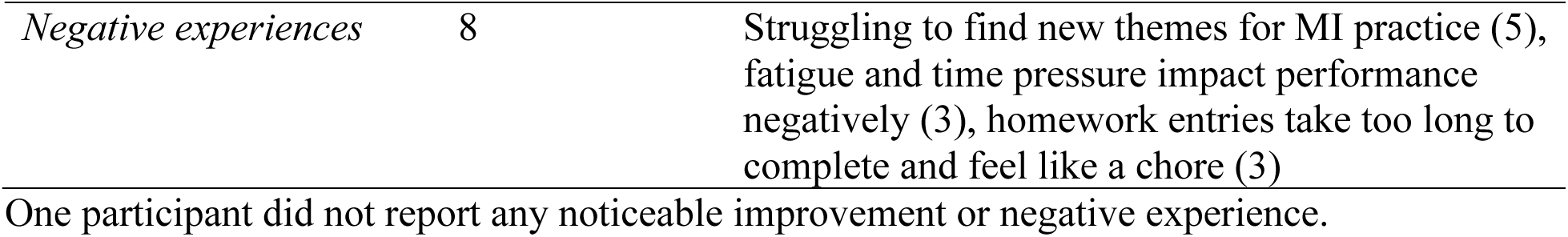
Experience with MI practice.

|  | # Participants | Examples |
| --- | --- | --- |
| *Improvements* |  |  |
| Planning and organizing | 14 | Helps plan and prepare for events, prioritize tasks, get organized, and structure the day |
| Self-regulation | 7 | Calming, relaxing, getting better at stress management, feeling more present, less anxious |
| Attention and concentration | 4 | Increased alertness and mental clarity, more engaged, better at staying on task, responding faster, more focused |
| Awareness | 4 | More aware of surroundings and sensations |
| Memory | 2 | Remembering items on the shopping list, old memories spontaneously emerge |
| *Negative experiences* | 8 | Struggling to find new themes for MI practice (5), fatigue and time pressure impact performance negatively (3), homework entries take too long to complete and feel like a chore (3) |

One participant did not report any noticeable improvement or negative experience.

**Table S5.** Head motion during scans.

|  | PD-MI | PD-Con | p |
| --- | --- | --- | --- |
| Mean (mm) | 0.57 ± 0.06 | 0.60 ± 0.07 | 0.214 |
| Maximum (mm) | 1.36 | 1.44 | - |
| # Invalid scans | 20.1 ± 15.4 | 25.9 ± 17.6 | 0.350 |

**Table S6.** MI themes practiced in the scanner.

| **Event planning** | **Household task** | **Outdoor task** | **Social Event** |
| --- | --- | --- | --- |
| Celebration (16) | Routine maintenance (17) | Grocery shopping (11) | Gathering (24) |
| Travel (10) | Cooking/preparing meal (7) | Gardening/yard work (8) | Special occasion (6) |
| Special event (3) | Home project (5) | Running errands (6) |  |
| Other (1) | Other (1) | Physical activity (5) |  |

The frequency of themes per category practiced by all subjects is shown. Examples of themes include:
Celebration: birthday, holiday, anniversary, party
Travel: Vacation, trips, visits
Special event: Wedding, funeral, religious
Routine maintenance: Cleaning, organizing, daily chores
Cooking/preparing meal: Breakfast, dinner, cooking for special occasion
Home project: Repairs, renovations, fixing things
Physical activity: Exercise, hiking, surfing, yoga
Running errands: Going to post office, cleaners, appointments

Scoring key:
| Improvement (+) or Decline (-) |  |  |  |  |  |
| --- | --- | --- | --- | --- | --- |
|  | None [0] | Slight [1] | Mild [2] | Moderate [3] | Significant [4] |
| Cognitive domain |  |  |  |  |  |

**Table S7.** Significant subjective changes in cognitive function at 6 weeks.

|  | MI (N = 10) | PE (N = 8) | p |
| --- | --- | --- | --- |
| Executive | 1.30 ± 1.14 (95% CI[0.49, 2.11]) | 0.29 ± 1.27 (95% CI[-0.77, 1.35]) | 0.027 |
| Memory | 0.57 ± 0.35 (95% CI[0.31, 0.82]) | 0.08 ± 0.56 (95% CI[-0.38, 0.55]) | 0.027 |
| Social cognition | 1.00 ± 0.94 (95% CI[0.33, 1.67]) | 0.00 ± 0.53 (95% CI[-0.45, 0.45]) | 0.021 |
| All cognitive domains averaged | 0.74 ± 0.33 (95% CI[0.50, 0.97]) | 0.14 ± 0.85 (95% CI[-0.57, 0.86]) | 0.006 |

Non-normally distributed
Nonparametric Mann-Whitney U test

